# Assessing Mpox knowledge, Attitude, and Willingness to accept the Mpox Vaccine among people living with HIV and men who have sex with men in Rivers State, Nigeria

**DOI:** 10.1101/2025.08.14.25333538

**Authors:** Chizaram Onyeaghala, Vivian Ifeoma Ogbonna, Ifeyinwa Philippa Ugboma, Nelson Oruh

**Affiliations:** Department of Internal Medicine, University of Port Harcourt Teaching Hospital, Port Harcourt, Rivers State, Nigeria; Department of Community Medicine, University of Port Harcourt Teaching Hospital, Port Harcourt, Rivers State, Nigeria; Initiative for Advancement of Humanity, Port Harcourt, Rivers State, Nigeria

**Keywords:** Mpox, PLHIV, MSM, vaccine hesitancy, knowledge, mpox vaccine, Nigeria

## Abstract

Limited data exist on knowledge and attitudes towards mpox and the willingness to accept the mpox vaccine among vulnerable populations, such as people living with HIV and men who have sex with men (MSM), in countries facing intersecting HIV and mpox epidemics, like Nigeria. This study assessed the knowledge, attitudes, and willingness of these groups to receive the mpox vaccine in Rivers State. A cross-sectional study was conducted from August 26, 2024, to September 30, 2024, involving 300 people living with HIV (PLWH) and 14 MSM at two selected health facilities in Rivers State, Nigeria, using systematic and snowball sampling techniques, respectively. Data were collected via a self-administered, web-based Google form covering sociodemographic information, knowledge of mpox, attitudes towards the disease, and vaccine willingness. The chi-square test explored the relationships between sociodemographic factors and vaccine acceptance. Multivariate logistic regression identified determinants of vaccination willingness. Although most participants (72.7%; 221) were aware of mpox, 54.3% (165) demonstrated poor knowledge of the disease. While 60.5% (184) perceived mpox as a serious illness, 60.2% (182) expressed willingness to accept the vaccine; however, concerns about side effects and mistrust in health systems served as key barriers. Willingness to vaccinate was significantly associated with age (χ² = 9.781; p < 0.007) and knowledge of mpox (χ² = 7.272; p < 0.027). Additionally, sex (χ² = 16.19, p < 0.001), level of education completed (χ² = 37.63, p < 0.001), and marital status (χ² = 15.01, p < 0.001) showed significant associations with mpox knowledge. The study revealed a concerning level of poor knowledge, limited perceived risk, and suboptimal vaccine acceptance, despite high awareness among PLWH and MSM. Vaccine acceptance was higher among younger individuals and those with good knowledge of mpox. Targeted public health education and confidence-building strategies are crucial for enhancing vaccine uptake among these at-risk groups.

**Summary box:**

- Limited data exist on the knowledge, attitudes towards mpox, and willingness to accept the mpox vaccine among vulnerable populations, such as people living with HIV (PLHIV) and men who have sex with men (MSM) in countries with overlapping HIV and mpox epidemics, like Nigeria.
- Vaccination remains a vital public health tool for controlling infectious diseases, such as mpox, within communities. However, factors affecting vaccine uptake include individual awareness, cultural beliefs, trust in healthcare, and the stigma associated with vaccination. These elements are especially important in Nigeria, where PLHIV and MSM often face systemic discrimination in healthcare settings.
- Our study data reveal a concerning level of poor knowledge about mpox, low perceived risk, and suboptimal vaccine acceptance, despite high awareness of the disease among PLHIV and the MSM community.
- Vaccine acceptance was influenced by younger age and a good knowledge of mpox.
- Targeted public health education and confidence-building strategies are essential for increasing vaccine uptake among these at-risk groups.

## Introduction

Human mpox (hMPX) is an emerging zoonotic infection caused by the monkeypox virus (MPXV), a DNA (deoxyribonucleic acid) virus belonging to the *Orthopoxvirus* genus and the Poxviridae family. ^1,2^ Historically limited to parts of Central and West Africa, the disease became a global concern following the 2022 outbreak that mainly affected non-endemic countries. ^3^ Before the discovery of a new strain (clade 1b) during the ongoing 2024-25 outbreak in East and Central Africa, there were generally two main clades of MPXV (clade I, clade IIa, and IIb). Clade II usually causes milder clinical disease than clade I, which is associated with a more severe clinical course and higher mortality. ^4^ Mpox typically begins with systemic symptoms such as fever, chills, muscle pain, and back pain, followed by skin eruptions and lymphadenopathy. It usually resolves on its own, but can be severe in pregnant women, individuals with chickenpox coinfection, and people living with HIV (PLHIV). ^5–7^

The 2022-2023 global mpox outbreak mainly affected men who have sex with men (MSM) and highlighted the role of sexual transmission of MPXV in mpox spread. ^3^ Although studies from Nigeria did not include the MSM population because of stigma, discrimination, criminalization of same-sex relationships, and limited healthcare access, ^8,9^ MSM remain a vital but underrepresented group in the mpox disease landscape. ^10^ People living with HIV (PLHIV) are at higher risk for severe mpox due to their weakened immune systems and social factors. ^11^ Despite the increased vulnerability of these groups, knowledge and perceptions about mpox and its prevention remain limited.

Vaccination remains one of the most effective public health measures for controlling the spread of infectious diseases, such as mpox, in the community. ^12–14^ However, vaccine uptake is influenced by several factors, including individual knowledge, cultural beliefs, trust in healthcare, and stigma; issues that are especially important in Nigeria, where PLHIV and MSM often face systemic healthcare discrimination. ^15,16^ Because vaccine hesitancy (refusal or delay of vaccination despite available services) has become an increasingly significant global health threat during the COVID-19 pandemic and is listed by the WHO as one of the top ten health threats facing humanity, it has become crucial to explore willingness to get vaccinated to develop and implement targeted public health strategies to control the virus spread effectively. ^17,18^

As Rivers State continues to report new mpox cases, there is an urgent need to evaluate awareness levels, attitudes toward prevention, and willingness to receive the mpox vaccine among high-risk populations. Failing to address these knowledge gaps could hinder public health efforts in controlling the outbreak. This is especially important since Rivers State is one of the states selected for the upcoming second phase of mpox vaccine distribution targeting at-risk groups such as PLHIV and MSM.

This study aims to fill this gap by evaluating the knowledge, attitudes, and willingness to vaccinate among PLHIV and MSM in selected health facilities in Rivers State, Nigeria. The findings will support policymakers, public health professionals, and HIV program managers in creating effective mpox vaccination strategies. By addressing knowledge and perception gaps, the study aims to enhance vaccine confidence, reduce transmission, and ultimately protect those most vulnerable.

## Materials and methods

### Study area

The study was conducted at two selected health facilities-the University of Port Harcourt Teaching Hospital and the Initiative for Advancement of Humanity in Rivers State, Nigeria. Rivers State, located in the southern region of Nigeria, is one of the country’s oil-rich states and serves as a central hub for commerce and healthcare activities in the Niger Delta. The state has a diverse population, including urban and peri-urban communities, with a mix of ethnic and socio-economic groups. The tertiary hospital chosen for this study, the University of Port Harcourt Teaching Hospital, is a leading healthcare institution in Rivers State, known for its comprehensive medical services, including specialized care for people living with HIV (PLHIV). This hospital is a 950-bed facility providing tertiary, secondary, and primary healthcare services due to the near-collapse of other facilities in the region. ^19^ It operates an intense HIV/AIDS treatment and care program, offering antiretroviral therapy (ART), counselling, and other support services. Additionally, it functions as a referral center for various infectious diseases, including emerging zoonotic diseases such as mpox. The other facility, Initiative for Advancement of Humanity, serves a significant number of MSM, a high-risk group for mpox, making it an ideal location to assess knowledge, attitudes, and willingness to accept the mpox vaccine. The HIV clinic at the University of Port Harcourt Teaching Hospital in Rivers State has an estimated 5955 active clients, making it the largest in Nigeria’s South-South geopolitical zone. The study area also benefits from support groups working to improve awareness and access to healthcare among key populations. Rivers State’s public health infrastructure emphasizes disease surveillance and vaccination campaigns, making it relevant to explore vaccine acceptance within this context.

### Study design

A cross-sectional study was conducted to survey participants at the two selected health facilities in Rivers State.

### Study population

The study participants include people living with HIV (PLHIV) attending the HIV clinic at the University of Port Harcourt Teaching Hospital and MSM attending the Initiative for Advancement of Humanity (IAH) in Rivers State, Nigeria. Those who were too ill to respond to the questions and individuals under 18 years old were excluded.

### Sample size determination

We used Cochran’s formula for calculating the sample size for cross-sectional studies. ^20^ Since there was no existing data on the knowledge, attitude, and willingness of PLHIV to accept the Mpox vaccine in Nigeria, we relied on the proportion of monkeypox vaccination intention among high-risk populations from a previous study in Nigeria, which was 83%. ^21^ With Z = 1.96 at a 95% confidence level, a margin of error of 5%, and a 10% non-response rate, our minimum sample size was 241. We surveyed a total of 304 respondents.

### Sampling technique

A systematic sampling method was employed to recruit participants from the clinic. The first participant was selected through simple random sampling by ballot from August 26, 2024, to September 30, 2024. The HIV clinic sees about 200 to 250 clients each day. Respondents were recruited based on the study’s inclusion criteria until the required sample size was reached. For enrolling participants from the MSM population, a snowball sampling technique was employed.

### Data collection method

The data were gathered using a questionnaire developed based on a review of relevant literature on mpox and the mpox vaccine. The questionnaire was created with Google Forms. Google Forms is an electronic survey tool used to distribute surveys to study participants. It was a self-administered questionnaire. The questionnaire included the following sections: a. Knowledge of the disease, b. Perception of mpox, and c. Willingness to receive the mpox vaccine. All study data were de-identified and stored on password-protected electronic devices. The tool’s reliability and validity were confirmed through pre-testing (Cronbach’s alpha = 0.78) and expert review.

### Statistical analysis

The de-identified research data (Excel sheet) was transferred to the Statistical Package for Social Sciences, now known as Statistical Product and Service Solutions (SPSS) version 27 (IBM Corp., Armonk, NY, USA), for statistical analysis. Sociodemographic characteristics and participants’ responses to the questions were uniformly defined as categorized data and summarized using frequencies and proportions. A chi-square test was used to assess the relationship between the independent variables (sociodemographic information) and the outcome variables (participants’ knowledge of the mpox vaccine, attitude toward the mpox vaccine, and willingness to accept the mpox vaccine). Variables with p < 0.05 in the chi-square test were included in a multivariable logistic regression analysis to identify factors associated with participants’ willingness to be vaccinated against mpox. For those variables included in the multivariable logistic regression, adjusted odds ratios (ORs) and 95% confidence intervals (CIs) were reported, with statistical significance set at p < 0.05.

### Ethical approval

The study received approval from the University of Port Harcourt Teaching Hospital research and ethical committee (NHREC/UPTHREC/02/2024). All participants were informed about the purpose, procedures, and their rights. They were also told that their identities or information would remain confidential. Written informed consent was obtained from each participant before the commencement of the study. The research was conducted in accordance with the principles outlined in the Declaration of Helsinki.

## Results

As shown in Table 5, 183 (60.2%) are willing to accept the mpox vaccine.

**Table 1:**
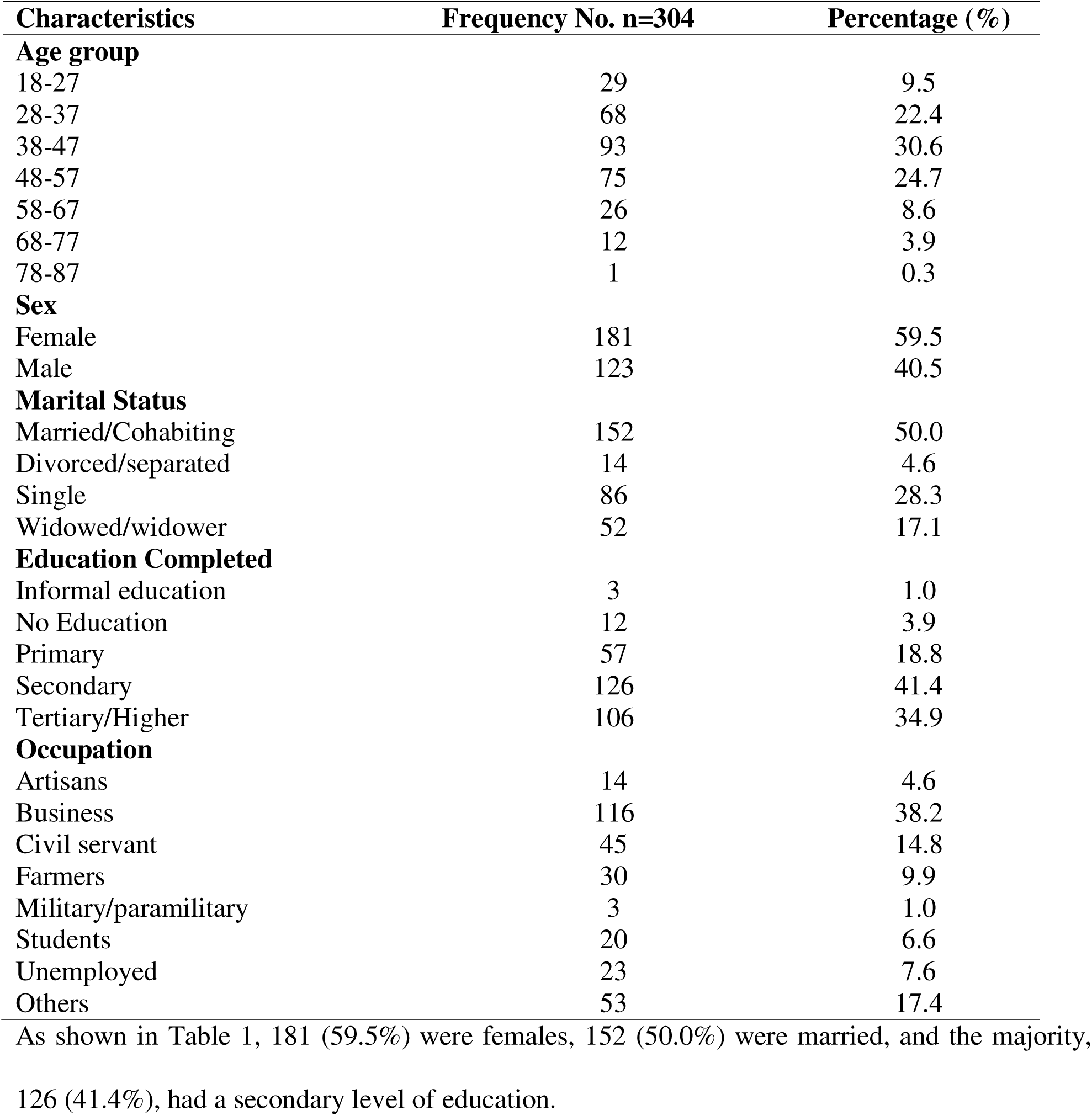
Socio-demographic characteristics of PLWH and MSM in selected health facilities in Rivers State, Nigeria.

| Characteristics | Frequency No. n=304 | Percentage (%) |
| --- | --- | --- |
| **Age group** |  |  |
| 18-27 | 29 | 9.5 |
| 28-37 | 68 | 22.4 |
| 38-47 | 93 | 30.6 |
| 48-57 | 75 | 24.7 |
| 58-67 | 26 | 8.6 |
| 68-77 | 12 | 3.9 |
| 78-87 | 1 | 0.3 |
| **Sex** |  |  |
| Female | 181 | 59.5 |
| Male | 123 | 40.5 |
| **Marital Status** |  |  |
| Married/Cohabiting | 152 | 50.0 |
| Divorced/separated | 14 | 4.6 |
| Single | 86 | 28.3 |
| Widowed/widower | 52 | 17.1 |
| **Education Completed** |  |  |
| Informal education | 3 | 1.0 |
| No Education | 12 | 3.9 |
| Primary | 57 | 18.8 |
| Secondary | 126 | 41.4 |
| Tertiary/Higher | 106 | 34.9 |
| **Occupation** |  |  |
| Artisans | 14 | 4.6 |
| Business | 116 | 38.2 |
| Civil servant | 45 | 14.8 |
| Farmers | 30 | 9.9 |
| Military/paramilitary | 3 | 1.0 |
| Students | 20 | 6.6 |
| Unemployed | 23 | 7.6 |
| Others | 53 | 17.4 |

**Table 2:**
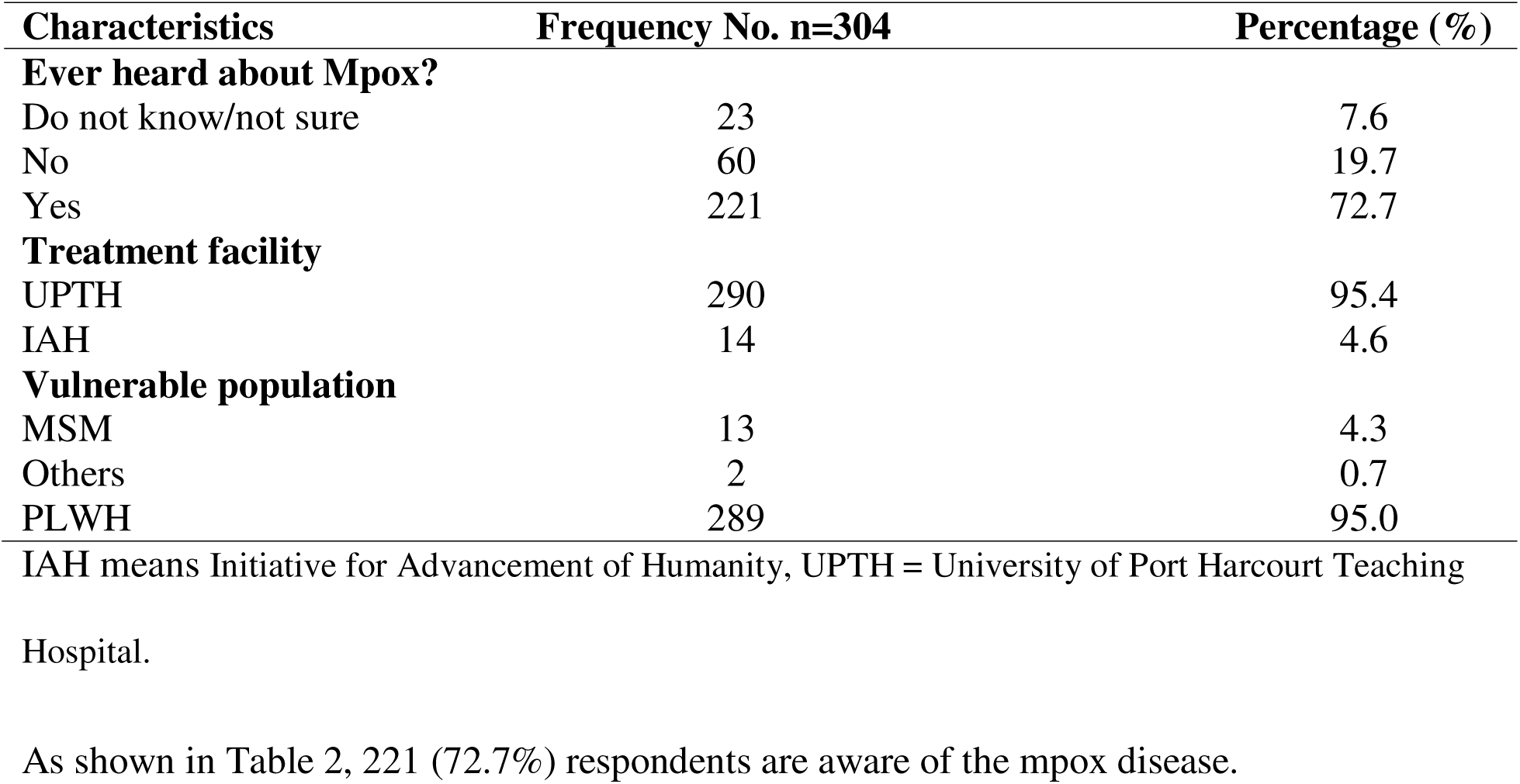
Awareness about Mpox among PLWH and MSM in selected health facilities in Rivers State, Nigeria.

| Characteristics | Frequency No. n=304 | Percentage (%) |
| --- | --- | --- |
| **Ever heard about Mpox?** |  |  |
| Do not know/not sure | 23 | 7.6 |
| No | 60 | 19.7 |
| Yes | 221 | 72.7 |
| **Treatment facility** |  |  |
| UPTH | 290 | 95.4 |
| IAH | 14 | 4.6 |
| **Vulnerable population** |  |  |
| MSM | 13 | 4.3 |
| Others | 2 | 0.7 |
| PLWH | 289 | 95.0 |

IAH means Initiative for Advancement of Humanity, UPTH = University of Port Harcourt Teaching Hospital.

**Table 3:** Knowledge of Mpox among PLWH and MSM in selected health facilities in Rivers State, Nigeria.

| **Characteristics** | **Frequency No. n=304** | **Percentage (%)** |
| --- | --- | --- |
| **Causes of mpox** |  |  |
| A bacterium | 14 | 4.6 |
| A Parasite | 2 | 0.7 |
| A virus | 109 | 35.9 |
| Do not know/not sure | 172 | 56.6 |
| Witchcraft/sorcery | 7 | 2.3 |
| **Fever is a symptom of mpox** |  |  |
| Do not know/not sure | 141 | 46.4 |
| No | 5 | 1.6 |
| Yes | 158 | 52.0 |
| **Chest pain is a symptom of mpox** |  |  |
| Do not know/not sure | 220 | 72.4 |
| No | 54 | 17.8 |
| Yes | 30 | 9.9 |
| **Body pain is a symptom of mpox** |  |  |
| Do not know/not sure | 172 | 56.6 |
| No | 28 | 9.2 |
| Yes | 104 | 34.2 |
| **Skin rash is a symptom of mpox** |  |  |
| Do not know/not sure | 105 | 34.5 |
| No | 5 | 1.6 |
| Yes | 194 | 63.8 |
| **Genital rash /ulcer is a symptom of mpox** |  |  |
| Do not know/not sure | 220 | 72.4 |
| No | 50 | 16.4 |
| Yes | 34 | 11.2 |
| **Dizziness is a symptom of mpox** |  |  |
| Do not know/not sure | 196 | 64.7 |
| No | 14 | 4.6 |
| Yes | 94 | 30.9 |
| **Overall knowledge score** |  |  |
| Poor (0-2) | 165 | 54.3 |
| Fair (3-4) | 59 | 19.4 |
| Good (5-7) | 80 | 26.3 |

**Table 4:** Risk Perception about the disease called Mpox among PLWH and MSM in selected health facilities in Rivers State, Nigeria.

| Characteristics | Frequency No. n=304 | Percentage (%) |
| --- | --- | --- |
| **Mpox is a severe disease.** |  |  |
| Do not know/not sure | 1 | 0.3 |
| Maybe | 96 | 31.6 |
| No | 23 | 7.6 |
| Yes | 184 | 60.5 |
| **At risk of getting mpox** |  |  |
| Do not know/not sure | 1 | 0.3 |
| Maybe | 66 | 21.7 |
| No | 161 | 53.0 |
| Yes | 76 | 25.0 |
| **Worried about getting mpox** |  |  |
| Do not know/not sure | 1 | 0.3 |
| Maybe | 45 | 14.8 |
| No | 177 | 58.2 |
| Yes | 81 | 26.6 |
| **Worried about transmitting mpox to your spouse/family** |  |  |
| Do not know/not sure | 2 | 0.7 |
| Maybe | 51 | 16.8 |
| No | 177 | 58.2 |
| Yes | 74 | 24.3 |

**Table 5:**
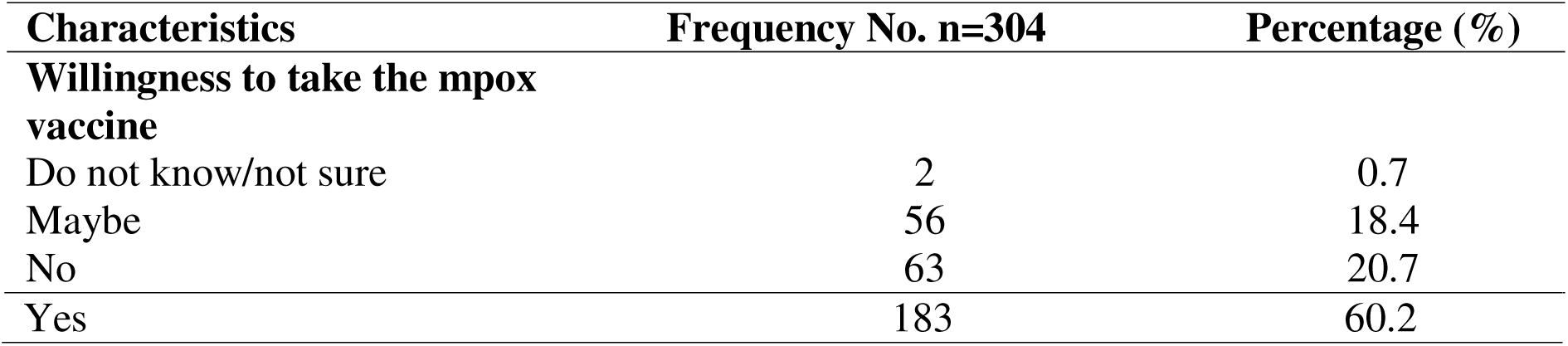
Willingness to accept the Mpox Vaccine in the study population.

As shown in Supplemental Table 6, the most common reasons for refusal to accept the mpox vaccine were that individuals did not think they could contract mpox, followed by uncertainty about the vaccine’s safety, and a lack of trust in the vaccine.

As shown in Supplemental Table 7, age and knowledge level were significantly associated with willingness to accept the mpox vaccine. Specifically, those ≤24 (youths) were more willing, 14 (100.0%), to receive the mpox vaccine, followed by the senior citizens (61-84 years), 152 (58.0%) (χ2 = 9.781; p < 0.007). Additionally, a higher proportion of those with good knowledge, 58 (72.5%) of mpox, were more willing to accept the vaccine compared to those with poor knowledge, 90 (54.6) (χ2 = 7.272; p < 0.027). Similarly, a higher proportion of those with a tertiary level of education (68, 64.2%) were willing to accept the mpox vaccine compared to those with no education (7, 58.3%) and informal education (1, 33.3%). However, this difference was not statistically significant.

As shown in supplemental table 8, sex, level of education completed, and marital status were significantly associated with knowledge of mpox. Other factors, such as age and occupation, were not statistically significant. Specifically, a higher proportion of males had good knowledge of mpox (35, 28.5%), compared to females (45, 24.9%) (χ2 = 16.19; p < 0.001). Furthermore, a higher proportion of those with a tertiary level of education (36, 34.0%) had good knowledge of mpox compared to those with no education (1, 8.3%) and informal education (0, 0.0%) (χ2 = 37.63; p < 0.001). Additionally, a higher proportion of those who are single had good knowledge of mpox 32 (37.2%) (χ2 = 15.01; p < 0.019).

## Discussion

To the best of our knowledge, this is the first comprehensive study to examine the level of knowledge and attitudes towards Mpox, as well as the willingness to receive the Mpox vaccine among PLWH and MSM in Nigeria. Available literature shows a lack of studies assessing mpox vaccination willingness in PLHIV and MSM populations. ^22–32^ Our study data reveal a concerning level of poor knowledge about Mpox, limited perceived risk, and subpar vaccine acceptance, despite relatively high awareness of the disease. These findings offer valuable insights into the behavioural and informational gaps that may affect vaccine uptake among these high-risk groups.

The finding that over half of the respondents lacked sufficient knowledge of the disease, with just over a quarter demonstrating a good understanding of mpox, highlights a serious public health concern that could hinder prevention efforts. Although 72.7% of participants were aware of mpox, awareness alone did not guarantee they had an accurate or thorough understanding of the disease. This knowledge gap is particularly troubling because PLHIV often attend HIV treatment and care clinics where they receive counselling and educational sessions, which might have boosted their awareness. The disconnect between awareness and understanding of mpox likely contributed to the spread of misinformation, delayed symptom recognition, and poor health-seeking behaviours during outbreaks, as noted in previous studies on Mpox in Nigeria and Cameroon. ^33,34^

These findings align with the results of Al-Mustapha and colleagues, who found that while a large proportion of Nigerians had heard of Mpox, only a small number could correctly identify transmission routes or prevention strategies. ^35^ The low knowledge level may be due to inadequate public education campaigns and Mpox being relatively less familiar compared to other common viral infections, such as HIV or COVID-19.

Despite the generally low level of knowledge among the study participants, 60% viewed mpox as a severe disease, which aligns with findings from previous studies indicating that perceptions of disease severity often coexist with limited factual understanding. ^36–38^ This suggests that how the study population views mpox’s severity might be shaped by other factors such as misinformation, cultural beliefs, and historical mistrust in public health efforts. It could also mean that public health campaigns are effectively emphasizing the potential seriousness of the disease. These low-risk perceptions could impede vaccine uptake and the adoption of safe behaviours, especially in a population that is already vulnerable due to underlying immunosuppression or risky sexual networks.

Willingness to accept the mpox vaccine was modest at 60.2%, similar to a study conducted among PLHW in northern Nigeria that reported a desire to receive the mpox vaccine in 64.4% of respondents. Low-risk perception, concerns about vaccine safety, and mistrust of authorities and pharmaceutical companies were cited as key barriers. ^11^ Other reasons for vaccine hesitancy reported by *Illiyasu et al*., in contrast to the initial study, included anxiety about vaccine-antiretroviral drug interactions, perceived protection from antiretroviral treatment, and the newness of the vaccine. Vaccine hesitancy, driven by low-risk perception, fear of serious adverse events, and mistrust of vaccines, has also been observed in Nigeria during the COVID-19 pandemic and is likely linked to broader structural mistrust in the healthcare system. ^39–41^

MSM often face criminalization, stigma, and healthcare discrimination, which could discourage engagement with health systems and vaccine access. These structural barriers must be addressed through policies that prioritize equity and inclusion. Community-based organizations (CBOs) that work with key populations can serve as trusted intermediaries to deliver accurate public health education and facilitate vaccine uptake in this under-recognized population.

Age, a key predictor of vaccine hesitancy, was found to have a significant link with willingness to accept the mpox vaccine in this study. Youths under 24 years old, followed by seniors (61-84 years old), were notably more likely to express a willingness to get the mpox vaccine. These results may suggest that other factors, such as age, may not be the primary influence on vaccine attitudes within the study population. ^42,43^ Similarly, participants with good mpox knowledge were significantly more likely to accept the vaccine. This highlights the importance of health literacy and targeted messaging, especially for younger, digitally connected populations who may be more receptive to risk communication. Previous research supports the idea that knowledge is a strong predictor of vaccine acceptance, as seen during the COVID-19 vaccine campaign in Africa. ^44,45^

Additionally, educational level played a key role in vaccine acceptance in this study. Although not statistically significant, 64.2% of those with a tertiary education were willing to accept the mpox vaccine, compared to participants with no formal or informal education. This highlights that informational gaps could potentially hinder vaccine acceptance. This finding aligns with a recent Nigerian study, which found that higher educational attainment is associated with better knowledge and increased vaccine acceptance. ^46^ Education can enhance understanding of disease mechanisms, foster trust in the healthcare system, and increase willingness to participate in public health initiatives. Therefore, vaccine communication strategies should address these educational disparities by providing clear, accessible information about mpox vaccines, especially in local languages and simplified formats. ^42^

Demographic analysis further revealed that males, individuals with tertiary education, and single participants were more likely to demonstrate good knowledge of mpox. This indicates a gender disparity in mpox awareness among PLWH and MSM, emphasizing the need for targeted vaccine communication strategies that include gender-sensitive approaches and focus on less-educated populations and those in marital relationships who might not see themselves as at risk, even if they are part of high-risk groups.

### Strengths and Limitations of the Study

This study possesses some strengths that underscore its relevance to the field of public health.

First, including the MSM population, a high-risk yet often understudied group due to their hesitation to seek medical care because of fear of stigma and victimization resulting from the criminalization of same-sex relationships in Nigeria, could provide valuable data to guide public health interventions. Second, the timing of this research was particularly strategic as it coincided with the preparatory stage of Nigeria’s first phase of deploying the mpox vaccine across seven states that targeted high-risk populations, including PLHIV. The findings of this study could offer crucial insights to support the deployment of the upcoming second phase of the mpox vaccine among at-risk groups in Nigeria. Ultimately, the use of both systematic and snowball sampling techniques enabled the collection of accurate data from these vulnerable and often underrepresented populations.

The study, however, has some limitations. First, the cross-sectional design used in our study provides a snapshot of data at a single point in time. While helpful in describing prevalence and associations, it does not allow for examining changes, such as knowledge of mpox over time, or establishing causal relationships between variables. Second, although our sampling method employed a systematic random approach, this may introduce sampling bias, as the findings may not fully represent the broader population of PLHIV in Rivers State or Nigeria. Similarly, using snowball non-random sampling to recruit a small number of MSM may not adequately reflect the wider community’s knowledge and attitudes, thus limiting generalizability. Nonetheless, the findings can serve as a foundation for future studies in the MSM population.

The small number of MSM enrolled in this study prevented disaggregating the data into MSM and PLWH groups.

Finally, the data collection relied on self-administered questionnaires, which could introduce response bias. Participants may provide socially desirable responses or inaccurately represent their knowledge and attitudes towards mpox, which could potentially affect the validity of our findings.

## Conclusion

The gap between disease awareness, actionable knowledge, and willingness to vaccinate underscores the need for comprehensive public health education among PLWH and MSM. Incorporating mpox education and vaccination services into existing HIV care programs can also enhance accessibility and foster trust among patients. These findings directly impact Nigeria’s national mpox preparedness and response plan, which should now focus on culturally sensitive education, using peer-led and trusted community networks, and addressing structural mistrust. Future policies should encourage inclusive, stigma-free access to vaccines, especially for MSM, who are often underrepresented in public health initiatives.

## Contributorship statement

Chizaram Onyeaghala conceived this study and wrote the initial draft. Vivian Ogbonna analyzed the data and wrote the results. Ifeyinwa Ugboma and Nelson Oruh obtained the data. All authors revised the final manuscript for publication.

## Ethical Approval

The Research and Ethics Committee of the University of Port Harcourt Teaching Hospital, Nigeria, approved this study.

