## Supplemental file for "Assessing Mpox knowledge, Attitude, and Willingness to accept the Mpox Vaccine among people living with HIV and men who have sex with men in Rivers State, Nigeria"

**Supplemental Material Section**

Table 6: **Reasons for unwillingness to accept** **the** **Mpox Vaccine**

| **Characteristics** | **Frequency No. n=304** | **Percentage (%)** |
| --- | --- | --- |
| I do not think I can get mpox | 76 | **25.0** |
| I do not trust the vaccine | 39 | 12.8 |
| I am not sure of the safety/side effects of the vaccine | 59 | 19.5 |
| I do not think it is a severe disease that needs a vaccine | 11 | 3.6 |
| Others (mostly no reason, fear) | 119 | 39.1 |

**Table 7: Socio-demographic and knowledge factors associated with Willingness to accept the** **Mpox Vaccine	`**

| **Variables** | **Willingness** **(Freq%) n=304** | | **Total** | | **Chi-square (P-value)** |
| --- | --- | --- | --- | --- | --- |
|  | **Not Willing**  **n=121** | **Willing**  **n=183** | |  |  |
| **Age** |  |  |  | | 9.781 (**0.007) *** |
| ≤24 | 0 (0.0) | 14 (100.0) | 14 (100.0) | |  |
| 25-60 | 110 (42.0) | 152 (58.0) | 262 (100.0) | |  |
| 61-84 | 11 (39.3) | 17 (60.7) | 28 (100.0) | |  |
| **Sex** |  |  |  | | 0.000 (1.000) |
| Female | 72 (39.8) | 109 (60.2) | 181 (100.0) | |  |
| Male | 49 (39.8) | 74 (60.2) | 123 (100.0) | |  |
| **Marital Status** |  |  |  | | 6.922 (0.075) |
| Married/Cohabiting | 61 (40.1) | 91(59.9) | 152 (100.0) | |  |
| Divorced/separated | 5(35.7) | 9 (64.3) | 14 (100.0) | |  |
| Single | 27 (31.4) | 59 (68.6) | 86 (100.0) | |  |
| Widowed/widower | 28 (53.8) | 24 (46.2) | 52 (100.0) | |  |
| **Education Completed** |  |  |  | | 2.028 (0.759) |
| Informal education | 2 (66.7) | 1 (33.3) | 3 (100.0) | |  |
| No Education | 5 (41.7) | 7 (58.3) | 12 (100.0) | |  |
| Primary | 25 (43.9) | 32 (56.1) | 57 (100.0) | |  |
| Secondary | 51 (40.5) | 75 (59.5) | 126 (100.0) | |  |
| Tertiary/Higher | 38 (35.8) | 68 (64.2) | 106 (100.0) | |  |
| **Knowledge of Mpox** |  |  |  | | **7.272 (0.027) *** |
| Poor | 75 (45.5) | 90 (54.5) | 165 (100.0) | |  |
| Fair | 24 (40.7) | 35 (59.3) | 59 (100.0) | |  |
| Good | 22 (27.5) | 58 (72.5) | 80 (100.0) | |  |
| **Vulnerable population** |  |  |  | | 1.54 (0.561) |
| MSM | 6 (46.2) | 7 (53.8) | 13 (100.0) | |  |
| Others | 0 (0.0) | 2(100.0) | 2(100.0) | |  |
| PLHIV | 115 (39.8) | 174 (60.2) | 289 (100.0) | |  |

**Table 8: Factors associated with knowledge of Mpox**

| **Variables** | **Knowledge (Freq% %)** | |  | | **Total n=304** | **Chi-square (P-value)** |
| --- | --- | --- | --- | --- | --- | --- |
|  | **Poor**  **n=165** | **Fair n=59** | | **Good**  **n=80** |  |  |
| **Sex** |  |  |  | |  | **16.19 (0.001) *** |
| Female | 113 (62.4) | 23 (12.7) | 45 (24.9) | | 181 (100.0) |  |
| Male | 52 (42.3) | 36 (29.3) | 35 (28.5) | | 123 (100.0) |  |
| **Education Completed** |  |  |  | |  | **37.63 (0.001) *** |
| Informal education | 2 (66.7) | 1 (33.3) | 0 (0.0) | | 3 (100.0) |  |
| No Education | 8 (66.7) | 3 (25.5) | 1 (8.3) | | 12 (100.0) |  |
| Primary | 40 (70.2) | 7 (12.3) | 10 (17.5) | | 57 (100.0) |  |
| Secondary | 80 (63.5) | 13 (10.3) | 33 (26.2) | | 126 (100.0) |  |
| Tertiary/Higher | 35 (33.0) | 35 (33.0) | 36 (34.0) | | 106 (100.0) |  |
| **Marital Status** |  |  |  | |  | **15.01 (0.019) *** |
| Married/Cohabiting | 92 (60.5) | 25 (16.4) | 35 (23.0) | | 152 (100.0) |  |
| Divorced/separated | 9 (64.3) | 1 (7.1) | 4 (28.6) | | 14 (100.0) |  |
| Single | 33 (38.4) | 21 (24.4) | 32 (37.2) | | 86 (100.0) |  |
| Widowed/widower | 31(59.6) | 12 (23.1) | 9 (17.3) | | 52 (100.0) |  |
